# Use of cfDNA and exovesicle-DNA for the molecular diagnosis of chronic Chagas disease

**DOI:** 10.1101/2023.02.24.23286400

**Authors:** Noelia Lozano, Mercedes Gomez Samblas, Eva Calabuig, María José Giménez Martí, Maria Dolores Gómez Ruiz, José Miguel Sahuquillo Arce, José Miguel Molina Moreno, M. Trelis, Antonio Osuna

## Abstract

Chagas disease is, among others, considered a neglected tropical disease. Given the magnitude of the human movements that have occurred in recent years from Central and South America to other countries, Chagas should now be considered a disease of worldwide distribution, in which the transmission of the parasite is restricted to transplacental transmission or blood or organ donations from infected people.

Parasite detection in chronically ill patients is restricted to serological tests that only determine previous contact and not the presence of the parasite, especially in those patients undergoing treatment evaluation or in newborns.

In this study, we evaluate the use of nucleic acids from both circulating serum exovesicles and cell-free DNA (cfDNA) from 448 serum samples from immunologically diagnosed chronic chagasic patients, which were re-evaluated by nested PCR on the amplicons resulting from amplification with kDNA-specific primers 121F-122R. Of the total number of samples selected, 50 were used to isolate and purify exovesicles from circulating serum and cell-free DNA (cfDNA).

When the nucleic acids thus purified were assayed as a template and amplified with primers 121F-122R and SAT, a percentage positivity of 100% was obtained for all positive samples assayed with the kDNA-specific primers and 96% when SAT primers were used. However, isolation of cfDNA for *T. cruzi* and amplification with SAT primers also showed 100% positivity. Hence, both samples can be used in those cases where it is necessary to demonstrate the active presence of the parasite.

## INTRODUCTION

*Trypanosoma cruzi* is a flagellated protozoan etiologically responsible for American trypanosomiasis or Chagas Disease (CD); a disease considered a zoonosis within the so-called neglected tropical diseases. In addition to humans, CD involves wild or domestic mammals. Hematophagous insects belonging to the Triatominae family (Reduviid) are responsible for CD transmission, acting as invertebrate hosts or vectors. The protozoa multiply in the vector°s midgut as epimastigote forms and finally reach the end of the intestine and rectal ampulla giving rise to the infective form or metacyclic trypomastigotes. With insect defecation after biting, these infective forms are deposited on the skin and they will access the mammalian host through the wound or through mucous membranes invading the nucleated cells. Then, they can multiply as amastigote forms. Parasites differentiate into trypomastigotes, which disrupt the host cell, invade intercellular spaces and penetrate neighboring cells or enter the bloodstream, disseminating the infection through the blood. Blood trypomastigote forms will wait to be ingested by a new vector. Non-vectorial human infection can occur by oral, transfusional (1,2), or vertical routes from mother to fetus (3). These last two routes are of epidemiological importance in areas without vectorial transmission.

Chagas disease is endemic in a total of 21 countries in the Americas, from the southern United States to Argentina and Chile (4,5), with about 10 million people infected, and an annual incidence of 50,000 to 200,000 infections (5). The disease, considered for many years to be restricted to Meso- and South America, has spread to other parts of the world due to migratory processes. Human-to-human cases have been described in the United States, Canada, European countries, Australia or Japan, areas with varying degrees of migration from endemic areas (6), turning Chagas into a public health problem in European countries such as Spain, Italy or Sweden, among others, where strict control measures have thus been taken in blood banks (7), gynecological services (8) and transplant surgeries (9–11).

The course of the disease has two marked phases: acute and chronic. Acute Chagas disease (ACD) occurs within the first eight weeks, immediately after the infection. It usually presents with a high number of parasites circulating in the blood. The chronic phase (CCD), which develops about 3 and 8 weeks after infection, can last for decades with no or mild symptomatology and a very low parasitemia level. It is estimated that only 30% of cases develop pathognomonic symptoms of the disease (12–14). Such symptoms include cardiac and/or gastrointestinal symptoms disorders (15). These chronic patients (CCD) constitute the greatest epidemiological risk for disease transmission in countries where infection by insect vectors does not occur.

Diagnostic methods vary from direct test, such as xenodiagnosis including the combination with molecular techniques, (16) to immunological techniques, in which different antigens (native or recombinant) are used (17,18). High variability in the sensitivity and specificity among techniques has been related to, not only to the type of laboratory technique or antigen used, but also to geographical differences in parasite strains, as well as genetic differences between human populations, which may contribute to discrepancies in serological tests (19). This has led to a series of recommendations by the Pan American Health Organization (PAHO) and national guidelines, (20–22) recommending the confirmatory use of two serological tests in parallel, with a sensitivity of at least 98% (23) for a correct diagnosis of the disease. Even so, when serological tests are inconclusive, some guidelines recommend molecular tests, such as PCR, the technique of choice for the confirmatory diagnosis of *T. cruzi* parasitemia (24). WHO recommends PCR as a confirmatory test after screening of blood donors, in the diagnosis of acute or congenital infection, or for therapeutic follow-up after diagnosis of acute infection (23). However, conventional PCR is unsuitable for chronic CD diagnosis due to low parasitemia and parasite DNA blood levels in the chronic phase, together with the different DNA extraction and purification systems, that bring PCR sensitivity to about 50-90%, whereas its specificity stays close to 100% (25). Due to the aforementioned drawbacks and despite its high specificity, PCR is recommended only when serological tests are inconclusive according to the recommendations of expert committees in different countries, such as Brazil, Chile, USA or Spain (20–22).

Therefore, in this paper we present different methodologies, different primers, the combination of nested PCR on non-visible PCR amplicons, the use of cell-free circulating DNA (cfDNA), and the extraction of DNA from exovesicles circulating in the serum (EVs-DNA) of chronic patients in order to try to solve diagnostic problems and facilitate the detection of CD in the hospital setting.

## MATERIAL AND METHODS

### Human blood samples

This study was conducted at the Microbiology and Parasitology department of Hospital Universitario y Politécnico La Fe (HUyP-La Fe), Valencia, Spain and at Parasitology department Institute of Biotecnology University of Granada Spain, from 2011 to 2020. CD serologically positive serum samples screened with the LiaisonXL murex chemiluminescence kit and tittered with the Trinity Biotech IFA kit were used. Samples (n 448) included patients with suspected disease or with manifest symptoms. They can be considered representative of the different groups of the population affected by CD, most of whom adults (n 313), mostly born in Bolivia, except for the children born in Spain from chagasic mothers (n 135). Some corresponded to patients who had received treatment for CD and who came for re-evaluation as follow-up, and patients with compromised immunity with post-transplant immunosuppression therapy and one of the patients was HIV seropositive. According to sex, 302 (67.4%) patients were female and 146 (32.6%) were male. All of the patients gave written informed consent before starting the study and all protocols were approved on March 4, 2011, the study was expanded to include the use to isolate and purify exovesicles from circulating serum and cell-free DNA (cfDNA) on September 26, 2018, with registration numbers 201102400000408 and 672/CEIH/2018 respectively, by the ethical committee of Granada University.

### DNA isolation method from peripheral blood

Peripheral blood samples (5 mL) were collected in Vacutainer vacuum blood tubes with EDTA and then subjected to a specific lysis pretreatment before proceeding to the DNA isolation methods. Pretreatment consisted of mixing 1:1 blood and denaturing lysis buffer (6M guanidine hydrochloride, 10 mM urea, 10 mM TRIS-HCL, 10 mM TRIS-HCL, 20% (v/v) Triton X-100; pH was adjusted to 4.4) through brief shaking. The samples were incubated at 70°C for 10 min. Before proceeding to the next steps, the samples were kept for at least 48 h at room temperature.

The automated purification protocol of the Maxwell Blood DNA Purification kit (Promega Biotech) was chosen according to the guidelines approved by HUyP-La Fe and was used following the manufacturer’s instructions. The initial volume for each of the blood sample was 400 µL of pretreated peripheral blood.

### Isolation of circulating cell-free parasite DNA (cfDNA) from serum

For the serum samples, 5 mL of blood were taken in the BD Vacutainer SST II Advance tubes (Reference. 366468). Once vortexed for 5 min, each tube was centrifuged at 1,500 x g for 10 min and approximately 2.0 mL of serum were collected to perform the different tests.

The samples used for the assay of this protocol were 4 serum pools and two individual serum samples from patients with digestive disorders. Two pools were formed by three patients with cardiac pathology each, and two pools formed by three patients with non-specific symptoms. (Table S1).

For the isolation of circulating DNA (cfDNA) of the parasite from the patient’s serum, the MagMAXTM cell-free DNA isolation kit (Thermo Fisher Scientific) was used and the manufacturer’s instructions were followed. First, the 100 µL serum pools were centrifuged at 1,600 x g 10 min at 4°C. Once the first centrifugation was completed, the supernatant was subjected to a second centrifugation at the same speed and time as the previous one in order to remove any cellular debris in the serum. Next, the proteins contained in the supernatant were digested for 20 min at 60°C with 2 µL of Proteinase k (20 μg/mL). To the digestion product, 150 µL of buffer (30mTris-HCl pH 8.0, 10mM EDTA, 1% SDS) were added together with 5 µL of magnetic bead solution. The resulting volume (255 µL) of the mixture was vortexed intensively for 10 min and then centrifuged at 14,000 x g for 10 s. The resulting pellet was washed twice, first with 500 µL of wash buffer and then with 500 µL of 80% ethanol, and subjected to centrifugation (20 s at 1,3000 x g). Finally, the pellet was dried and resuspended in 50 µL Milli-Q water.

### DNA isolation from serum exovesicles (EVs-DNA)

To improve the detection of parasite DNA in *T. cruzi* infected patients, DNA was isolated from exovesicles (EVs) present in serum. For this assay, 25 nested PCR negative and 25 nested PCR positive samples were selected as described below.

Purification of circulating EVs from serum was performed following the methods previously described by Diaz et al. (26) and Retana et al. (27), by a mixed ultracentrifugation-ultrafiltration procedure. For this purpose, 1 mL serum samples were each diluted (1:1) with PBS previously ultrafiltered through 0.22 µm pore filters. The diluted samples were first centrifuged at 3,500 x g for 10 minutes (4°C) to eliminate contamination by cells or cellular debris from the serum. The pellet obtained was discarded and the supernatant was ultrafiltered through sterile 0.45 μm pore filters (Millipore, USA) in order to remove apoptotic debris and particles remaining in the supernatant from the first centrifugation. It was subsequently ultracentrifuged in microtubes (Hitachi No 1508) at 110,000 × g for 2 h at 4°C in a Sorwal WX80 centrifuge with fixed-angle rotor (Fiberlite™ F50L-24 × 1.5). The resulting pellet was washed three times by ultracentrifugation in sterile PBS as described above and evaluated by transmission electron microscopy and NTA nanoparticle tracking analysis, as described in a previous paper (28).

For the isolation of DNA from EVs-DNA, the method described by Orrego et al. in 2020 was followed. To do so, the sediment containing EVs from serum was treated with a total of 2 units of DNAase I at 37°C for 30 min to remove external DNA from the EVs. After treatment, the enzyme was inactivated with heat at 70°C in a solution containing 50 mM EDTA. After removal of free nucleic acids from the EVs suspension, 100 µL of EVs were lysed with 200 µL of lysis buffer (30 mMTris-HCl pH 8.0, 10 mM EDTA, 1% SDS) supplemented with 20 µL of proteinase K (0.1 mg/mL), shaken by vigorous pipetting and incubated at 56°C for 1 h. Finally, nucleic acid purification and precipitation were performed following the traditional phenol-chloroform method by mixing 320 µL of lysed EVs suspension with 320 µL of phenol-chloroform-isoamyl reagent, 25:24:1 (v/v) (Sigma). The mixture was shaken briefly and centrifuged at 14,000 x g for 10 min. The aqueous phase was then collected and the DNA was precipitated with 1/10 vol of 3M sodium acetate pH 5 and 2.5 vol of HPLC grade absolute ethanol. The mixture was incubated overnight at -20°C and centrifuged at 15,000 x g for 10 min at 4°C, the supernatant was discarded and the precipitate was washed twice with cold 70% ethanol. Finally, the pellet was dried in a Jouan Thermo speed Vac (RC1010) at 20°C and diluted in 20 µl Milli-Q water.

### Transmission electron microscopy

To corroborate by electron microscopy the purification of EVs from serum, the ultracentrifuged button resulting from the purification process described above was fixed for 2h at room temperature in 2.5% glutaraldehyde in cacodylate buffer pH 7.2 and then resuspended in PBS for sample processing by negative staining. For this purpose, approximately 15 μL of the suspension were deposited on nickel 300 mesh grids, coated with a charcoal layer, for 10 min. The grids were then washed twice with ultrapure water for 1 min. The grids were negatively stained with 1% uranyl acetate for 1 min. After staining, the samples were dried on filter paper and observed under a Zeiss Libra 120 Plus transmission electron microscope at 120KV at the Centro de Instrumentación Científica (University of Granada facilities).

### Detection of *T. cruzi* DNA in biological samples by different PCR strategies Genes used in the study

In this work, two specific, conserved and highly repeated genes in the *T. cruzi* genome, widely used in the diagnosis of Chagas disease, were analyzed: the kinetoplast DNA (kDNA) minirepeat regions, which represent the major sequence component of kDNA with about 120,000 copies per parasite (29,30), and the nuclear satellite DNA with 10^4^ to 10^5^ copies in the highly conserved parasite genome (31).

### PCR strategies

#### The kDNA minirepeat regions and nested PCR

DNA from the 448 serologically positive samples, extracted using the Maxwell Blood DNA Purification Kit automated protocol (Promega Biotech), and the minirepeated region of the kDNA was amplified. The reaction mixture was made up of Go Taq1flexi 1X amplification buffer (25 mM dNTPs, 2.5 mM MgCl_2_, 1.0 U of Go Taq1(Promega, USA), 10 pmol of specific primers 121F and 122R (Table 1) and 10 µL of template. The final volume was 50 µL according to the methodology described previously (32). Amplification conditions consisted of two cycles, each of 1 min at 98°C and 2 min at 64°C, followed by 33 cycles of 1 min at 94°C, and 1 min at 64°C, before a final extension at 72°C for 10 min. In all cases, a DNA sample of *T. cruzi* from the culture as a positive control and a non-template control were used.

**Table 1.**
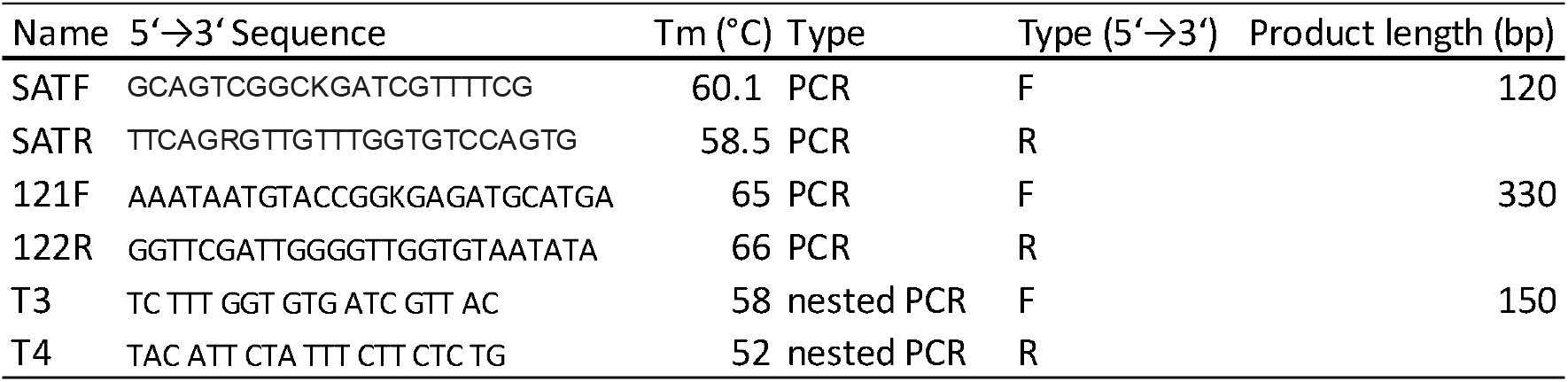
Sequences of primers used in this study.

In all instances, the amplification result, although not visible on DNA electrophoresis, was subjected to nested PCR amplification. To this end, 2 µL of PCR product were used as template, 0.2 µM concentration of T3 and T4 primers (Table 1), 0.2 mM of each dNTP, 3 mM MgCL_2_, 0.5 µL of GoTaq® Flexi DNA Polymerase and the final volume was 50 µL. The nested PCR conditions were (1) 94°C 5 min, (2) (94°C 1 min, 48°C 1 min, 72°C 1 min) × 35, (3) 72°C 7 min, (4) 12°C indefinite.

Amplifications were performed on a GeneAmp PCR System 9700 thermal cycler, AB Applied Biosystems and the nested PCR amplification product was sequenced on a GenomeLab GeXP system. The results obtained from sequencing were aligned using BLAST programs and the identity and coverage of these sequences were studied.

#### EVs-DNA and cfDNA PCR

Both the EVs-DNA present in the serum and the cfDNA were amplified indistinctly with the primer pair 121F-122R corresponding to the kDNA and with the SATF and SATR primers for the nuclear satellite regions (Table 1) using for these amplifications 0, 5 µM of the primers, 0.2 mM of each dNTP, 2.5 mM MgCl_2,_ 0.005 µL DMSO, 0.125 µL GoTaq Flexi DNA Polymerase enzyme and 2 µL as DNA template from DNA purification, the final reaction volume was 25 µL. For the SAT primers, the protocol was (1) 95°C 5 min, (2) (94°C 10 sec, 65°C 10 sec, 72°C 10 sec) × 40, (3) 72°C 5 min, (4). Samples were kept at 12°C.

The final products were visualized on a 2% agarose gel stained with Safe Sybr and developed on a ChemiDoc MP Imaging System (Biorad). Electrophoresis was performed at 100 mV for 30 min.

## RESULTS

### Amplification of DNA from peripheral blood

The sera of 448 immunologically positive subjects, as indicated above, came from patients who had visited the different medical services of the Hospital La Fe; 67.4% corresponded to samples from female patients and 146 samples to males (32.6%). The majority (313) of whom were born in American countries, mostly in Bolivia. While 135, namely the youngest, were born in Spain but of Latin American mothers.

Figure 1A shows the results of the first PCR with amplicons of 330 bp. Nested PCR (corresponding to the kDNA) gave a positive result for 72 (16%) samples in which the 150 bp amplicon was visible in agarose electrophoresis (Figure 1 B). Sequencing results of this amplicon are shown in Figure 1 C. The results of the NCBI BLAST analysis of these sequences verified that the amplicon corresponded to a kDNA region of *T. cruzi*, 150 bp with coverage of 93% and an E of 2e-44 and an identity percentage of 97.4% (Figure 1 C).

**Figure 1.**
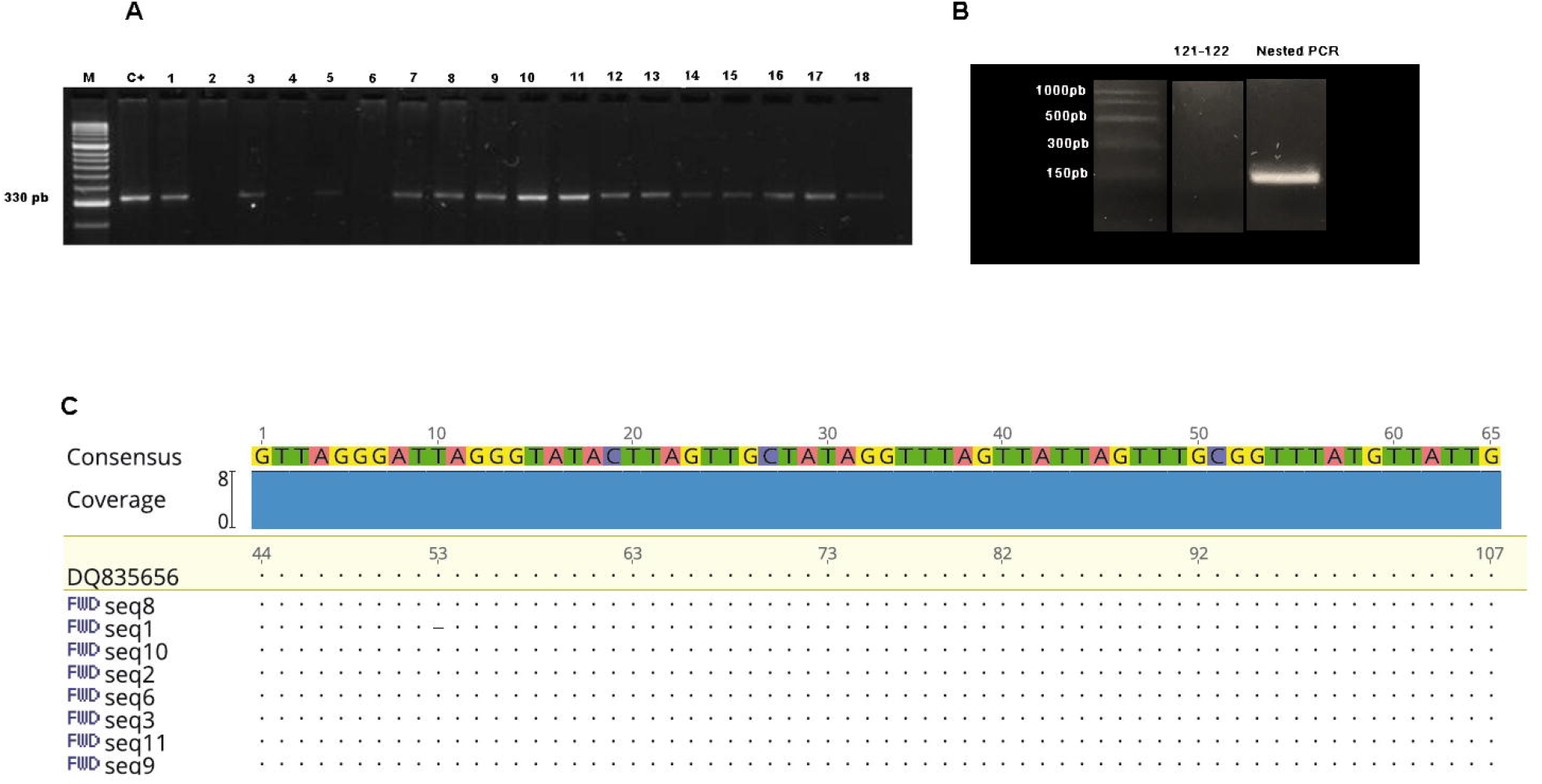
A: 2% agarose gel analysis of PCR-amplified products showing a 330 pb with 121 and 122 primers. PCR performed with peripheral blood DNA. Line 1, Hipper ladder II, Line 2, positive control, line 3 to 20 samples from 1 to 18 patients consecutively. B: 2% agarose gel showing a 150 pb band which belongs to nested PCR performed with T3 and T4 primers. C: Sequence alignment using as reference sequence the kDNA *T. cruzi* sequence with accession number DQ835656.

Analysis of the 72 samples, in which sample positivity was verified by nested PCR, resulted in 49 patients being female and 23 males. Mean age was 36.6 years (35 median) for females and 35.7 years (34 median) for males. Furthermore, positive patients were classified into 4 age ranges as shown in Table S2. Thirty-four (5.6%) women were of childbearing age and 17 of whom were pregnant. Also, *T. cruzi* DNA was detected in the peripheral blood sample in the three children who were between 0 to 2 years old.

Most of CD patients were born in Bolivia (n 70), one patient was from El Salvador and another one from Ecuador. Table S3 shows the distribution of origin in Bolivia, with Santa Cruz being the most represented region with 44.2% of the patients, followed by Sucre with 12.8%, Cochabamba with 10%, Potosí with 5.7%, La Paz (1.4%), Tarija (1.4%) and unidentified regions 25.2%.

Table S1 shows the conditions and age corresponding to the 72 positive nested PCR patients used in the study. Most of whom presented indeterminate symptoms (n 53), two had digestive disorders and nine patients suffered heart conditions. Of the total 72 samples, 25 corresponded to patients who had not yet been treated with benznidazole, and it is noteworthy that of the 47 that had been treated only three remained PCR positive for *T. cruzi*.

### DNA serum EVs and cfDNA amplification

#### Confirmation of circulating serum EVs

TEM visualization of EVs from serums pool isolated by ultracentrifugation is shown in Figure 2A. The mean size of the EVs was 119.92 ±18.41 nm. NTA analysis (Figure 2 B) revealed the different populations of these EVs in which the majority peak revealed a size of 163 nm. A second population less numerous of EVs with an average size of 231 nm was observed. Analysis of the total population of EVs showed a mode size of 208±84.8 nm and the mode was 163 nm. Also, few populations of larger size were observed which could be considered aggregates of the EVs purified from the circulating EVs in the serum of the patients studied.

**Figure 2.**
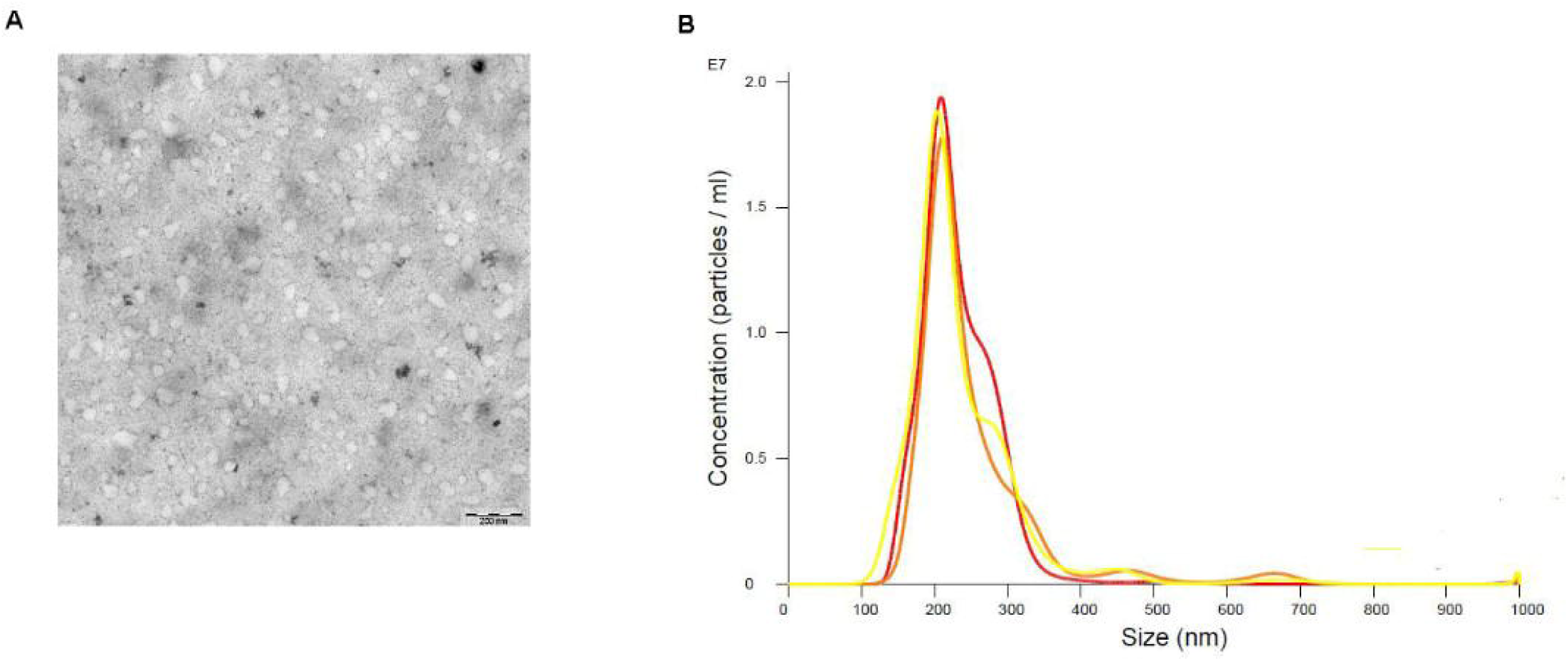
EVs characterization. A: isolated serum pool EVs observed under TEM. B; NTA showing the EVs populations.

#### Liquid biopsy

A total of 50 sera selected from the initial 448 serologically positive patients were used for these experiments. Twenty-five of these corresponded to patients who were not positive by nested PCR, while the remaining 25 sera were from patients positive by nested PCR. Serum EV DNA (EVs-DNA) and circulating free cellular DNA (cfDNA) were purified from the selected samples.

To perform PCR and as indicated in the Material and methods section, primers 121F-122R and those corresponding to SAT satellite DNA were used indistinctly with the DNA purified by these methods.

#### PCR analysis using DNA-EVs

The results of parasite EVs-DNA circulating in serum showed that twenty-five patients were positive for primers 121F-122R (Figure 3 A) and all but one (24) were positive for SAT primers (Figure 3 B).

**Figure 3.**
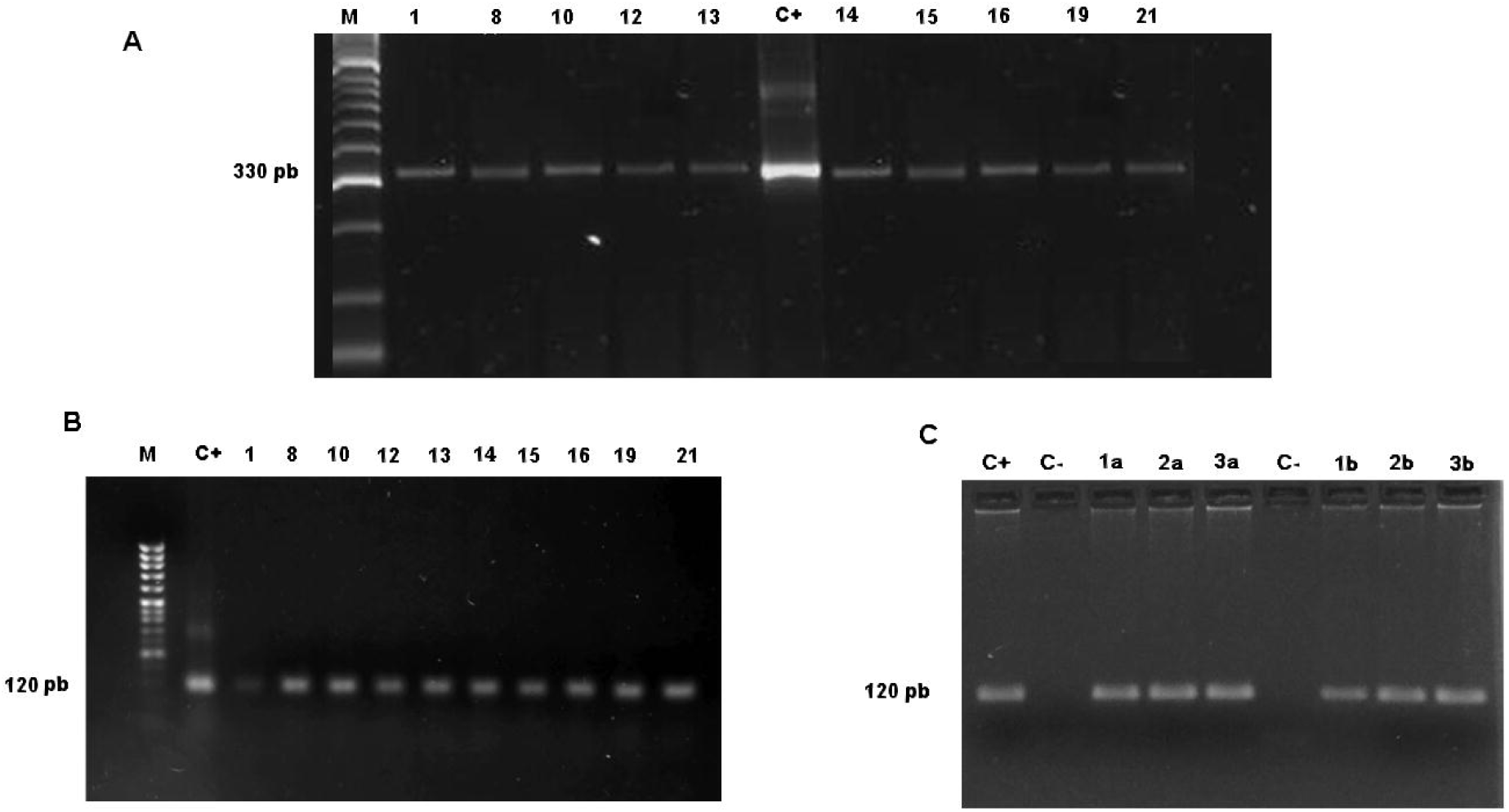
2% agarose gel electrophoresis analysis of PCR-amplified products. A: PCR performed with 121 and 122 primers and EVs-DNA. Line M, Hipper ladder II; Line 6, C+, positive control; line 2 to 5 and lines 7 to 11 samples of patients. B: PCR performed with SAT primers and EVs-DNA. Line M, Marker, Line 2, positive control; line 3 to 10 samples 1, 8, 10, 12, 13, 14, 15, 16, 19, 21 respectively. C: PCR performed with SAT primers and cfDNA. Line C+, positive control; Line C-non-template control; Line 3 to 5 samples 1a, 2a, 3a; line 6 non-template control; line 7 to 9 samples 1b, 2b, 3b.

#### PCR analysis using cfDNA

Isolated cfDNA was amplified by PCR with SAT primers. Figure 3 C shows six positive samples (Table S1) corresponding to chagasic patients with cardiac, digestive and indeterminate conditions. The same samples were negative when amplified by PCR with primers 121F-122R.

## DISCUSSION

In the last decades, the distribution of CD has spread due to human migration for economic reasons from geographical areas of Central and South America, where hematophagous vector transmission is still the case, to non-endemic areas of North America, Europe, Japan or Australia. Spain, Italy and Switzerland are the three European countries (33) with highest levels of Latin American communities, especially Spain, related to their common language as well as historical and cultural links. The health authorities in Spain and Tuscany (Italy), areas with a high level of Latin American migrants have therefore adopted standards and recommendations on hemodonations (34), and transplant donors with regard to the detection of chagasic patients in order to prevent human-to-human transmission of the disease (2,35). Similarly, regulations have been adopted for the screening of pregnant women originating from endemic countries and for which both the mother’s and newborn’s blood are tested (36,37) in order to establish a treatment, which is well tolerated and effective in this type of patient. All this has prompted the standardization of diagnostic methods for CD in health centers and especially for chronically ill patients, in whom parasitemia levels are practically non-existent.

Classical parasitological diagnostic methods, such as visualization of trypomastigote forms in blood smears or thick drop, or even trypomastigote concentration techniques by centrifugation in microhematocrit tubes, have been used to diagnose CD (38) (39). They show high specificity but very low sensitivity, especially in chronic patients. These methods may be applicable in diagnosis when the disease is developed with high parasitemia in the acute phase or after reactivation of circulating parasitemia in immunosuppressed chronic patients (38). Only culture and xenodiagnosis can be used to directly detect *T. cruzi* infections in the chronic phase. But these techniques are not useful in clinical practice, especially in Europe, due to the limitations of insect vector handling and the delay in obtaining results, which limits their usefulness in diagnostic studies and in evaluating treatment efficacy (40,41). Moreover, the evaluation of different techniques shows that in the case of parasitemia, PCR is at least 27 times more sensitive than blood culture (42).

The course of the disease is characterized by elevated IgM levels during the initial acute phase of the disease and the appearance of IgGs that persist throughout the disease (43). Thus, immunological diagnostic techniques are the gold standard for the diagnosis of CD in chronic patients, even in asymptomatic cases. The limiting factors in serological studies are the possible cross-reactivity with *Leishmania* spp. or *Trypanosoma rangeli* infections in those geographical areas where the disease was acquired and these trypanosomatids coexist (40,44). Another drawback could be the lack of sensitivity to the different parasite strains that affect humans throughout the Americas. They may present antigenic variability, which requires, if possible, the use of antigens of different geographical origin and different nature (19,45) according to the recommendations of the different committees on CD. Individuals are considered infected when they have a positive result in at least two serological techniques using different antigens. A third technique is required in case of discrepancy of indeterminate immunological results (20–23), or the use of a mixture of antigens that minimize discrepancies (17,46). The other significant case that invalidates immunological techniques is the diagnosis in newborns from infected mothers due to the presence of IgGs from the mother to the fetus by transplacental transfer (42,47–49).

In cases that require the detection of the direct presence of the parasite in blood, when doubtful or inconsistent serological results are obtained and when the efficacy of the patient’s treatment is evaluated, PCR methods capable of detecting the presence of parasite nucleic acids in blood (50) are needed to determine the unequivocal presence of the parasite. However, this technique, has a number of limitations that can alter the results and that must be considered (51,52) such us the volume of blood processed, the correct handling of the blood (use of the chaotropic agent guanidine, time and treatment of the lysis process), the manual or automatic DNA extraction procedures, the presence and especially the elimination of PCR inhibitors, the primers used, etc. For all of these reasons, the quest for new procedures and methodologies in the extraction of nucleic acids to be applied to minimize false negative results continues.

The samples reflect the Latin American population living and working in the area of influence of the Hospital La Fe in Valencia. Data from INE (National Institute of Statistics) for the year 2021 show that 17,847 people born in Bolivia reside in the Valencian Community, 39.1% are male and 60.9% are female.

A nested PCR was performed on all the samples, in which the primers T3 and T4 designed for this purpose were used and which would amplify a band of 150 bp from the internal zone of the amplicon produced in the first PCR. The use of nested PCR using other genes with the products of the first PCR was already studied by Pereira et al. (53) and Ribeiro et al. (54), who concluded that the sensitivity of a series of 3 nested PCRs using primers specific for conserved regions of the Kinetoplast is able to detect 1 parasite per ml of blood, increasing the sensitivity of the PCR by about 10,000 times. Other authors such as Marcon et al., 2002 (26), or Riera et al., 2006 (55) employed nested PCR after conventional PCR using TCZ1 and TCZ2 and internal primers TCZ3 and TCZ4 to evidence parasitization in a neonate. Also, Pereira et al., 2016 (53) used nested PCR for the products of a PCR using the TCZ1 and TCZ2 primers previously mentioned to detect CD positivity in a population of elderly people suffering from cardiac problems and in whom the involvement of *T. cruzi* in these pathologies was not suspected.

Sixteen percent of the total 448 immunologically positive patients tested positive in nested PCR.

The amplicons obtained were visualized by electrophoresis resulting in a band of 150 bp. Sequencing of the amplicons showed that they correspond to an internal region of the *T. cruzi* kDNA described by Burgos et al. (56), with a coverage of 93% and a percentage of identity of 97.4%, Figure (1).

The treatment of choice usually prescribed for CD-positive patients in Spain consists of 5 mg/kg benznidazole (BZ; N-benzyl-2-nitroimidazole acetamide) daily for 60 days. It is noteworthy that, of the 47 selected patients who had been treated, three tested positive post-treatment in nested PCR for *T. cruzi*. It was not ruled out that the treatment was not completed as a consequence of the side effects involved and the consequent interruption by the patient, and in one case the patient stated that he had returned to his country, not ruling out reinfection.

We also evaluated whether cfDNA present in the serum of CCD patients could be used as a template for specific PCRs to improve diagnosis in CCD and in neonates where parasitaemia is very low and almost undetectable by other diagnostic procedures. The results of the analysis were 100% positive. This confirms that serum is a good sample to carry out DNA extraction to be used in the PCR detection of *T. cruzi*, which was already determined by Russomando et al. (57) and in the review carried out by Weerakoon et al. (58) where the use of cfDNA in different biological samples in the diagnosis of different parasitoses was encouraged.

The results obtained in our cfDNA (nuclear DNA or mitochondrial KDNA) of parasitic origin would come from circulating parasites coming from either lysis or apoptosis of free or intracellular parasitic forms, or transported in elements secreted by the parasitic forms. With these sera we proceeded to the purification of EVs circulating in the serum, and the purification of the EVs from the serums of the patients was evidenced both by electron microscopy and by NTA. The term EVs designates the set of various EVs released by cells and delimited by a lipid bilayer that are released into the extracellular space by prokaryotic and eukaryotic cells (59). EVs constitute cellular mechanisms of cell-to-cell communication over short or long distances between cells acting as endocrine, paracrine, juxtacrine or autocrine signaling (27), through uptake of EVs or receptor-mediated interactions. Evidently, blood is the mechanism of transport and diffusion of these EVs. EVs are classified by their size, biogenesis and composition, exosomes with sizes between 20 to 200 nm, ectosomes between 200 to 1000 nm and apoptotic vesicles of a larger size (60). Exosomes contain functional molecules (including proteins, nucleic acids and lipids) derived from their cells of origin, being present in all biological fluids where they have been sought, urine and saliva and undoubtedly in blood (61–63). Due to the protection of the lipid bilayer, exosomes are relatively stable and proteins and other molecules transported in these small vesicles are protected from degradation. Their composition is quite complex, including a wide variety of lipids, proteins, different populations of RNAs, ssDNA, and metabolites (28). Typically, the internal volume of an exosome ranges from 20 to 90 nm^3^, suggesting that a prototypical exosome would contain approximately 100 proteins and 10,000 nucleotides (64).

Two PCRs were performed with the DNA extracted from the serum EVs, one with the primers (121F-122R) and the other PCR with the SATF and SATR primers; with the former, 50% positivity was obtained and with the SATF and SATR primers, 48% of the patients were positive.

The presence of parasite EVs in the serum of patients with CD was already described by some of us in 2017 (26) and these EVs appeared circulating in the serum forming immunocomplexes in patients with different Chagas pathologies, which denoted the presence of metabolically active forms of the parasite and capable of actively releasing these EVs.

EVs have been employed as liquid biopsy in numerous assays for tumor diagnosis (65–67), however the use of EVs-DNA for the diagnosis of pathogens and especially intracellular pathogens has hardly been addressed so far. Cho et al., 2020 (68) showed that PCR using exosome DNA from isolates of patients affected by tuberculosis shows higher sensitivity than conventional PCR diagnosis using total DNA for PCR. To our knowledge, this is the first study to report the detection of *T. cruzi* DNA from circulating EVs in the serum of patients using two sets of primers, i.e., those that detect kinetoplast DNA minicircles (121F-122R) and those that recognize nuclear DNA with the SATF and SATR primers.

From the results of the present work, it is evident that both cfDNA and EVs-DNA from the serum of infected patients constitute useful and effective biological samples to demonstrate, by PCR using conventional primers, the active presence of the parasite, in clinical cases (neonates and studies of the efficacy of treatment) where it is necessary to demonstrate the presence of the parasite.

## Data Availability

All data produced in the present work are contained in the manuscript in supplementary material

## FUNDING AND ACKNOWLEDGMENTS

This research was funded by the ERANet program, Research in prevention of congenital Chagas disease: parasitological, placental and immunological markers (ERANet17/HLH-0142 (Cochaco). Instituto Carlos III, Ministerio de Sanidad, Gobierno de España; Fundación Ramón Areces “Interactoma de las exovesículas de *T. cruzi* y de los inmunocomplejos que forman con las células del hospedador: implicaciones en la patología de la enfermedad de Chagas (2019)”. Ministerio de Ciencia y Tecnología of the government of Spain funded the project PGC2018-099424-B-I00 and The financial support given by the proyect A-BIO-350-UGR18 I+D+i Proyect “Programa Operativo FEDER de Andalucía JJAA” 2014-2020.

## SUPPORTING INFORMATION CAPTIONS

Table S1. Patients information

Table S2. Patients age ranges

Table S3. Distribution of origin in Bolivia in regions

