## Supplementary figures and images for "Use of cfDNA and exovesicle-DNA for the molecular diagnosis of chronic Chagas disease"

### Tables S2 and S3

Table S2. Patients age ranges

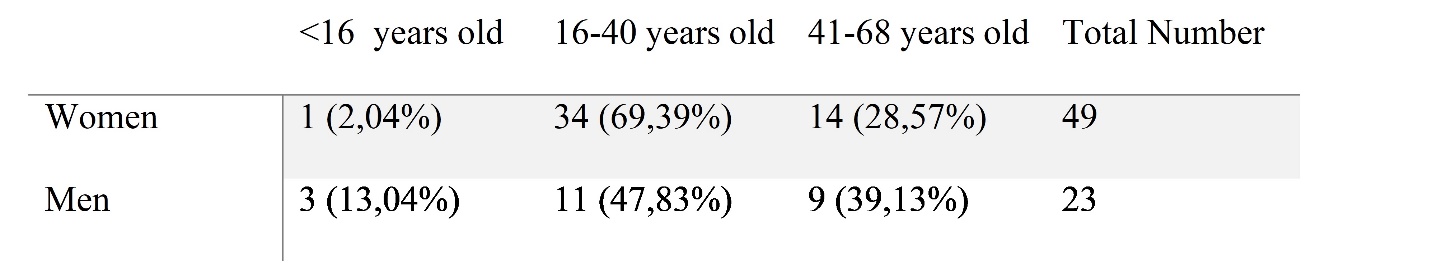

Table S3. Distribution of origin in Bolivia in regions

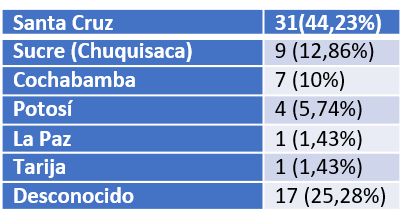
